# Evaluating Long-Term Autonomic Dysfunction and Functional Impacts of Long COVID: A Follow-Up Study

**DOI:** 10.1101/2024.10.11.24315277

**Authors:** Ella F. Eastin, Jannika V. Machnik, Nicholas W. Larsen, Jordan Seliger, Linda N. Geng, Hector Bonilla, Phillip C. Yang, Lauren E. Stiles, Mitchell G. Miglis

## Abstract

**Background:** The longitudinal prevalence and autonomic symptom burden in Long COVID patients is not well-established.

**Objective:** Assess the duration and severity of autonomic dysfunction in adults with Long COVID and evaluate its impact on function and quality of life.

**Design:** A follow-up survey of a subset of participants from a cross-sectional online survey of adults with Long COVID. Multivariable logistic regression identified predictors of moderate to severe autonomic dysfunction.

**Participants:** 526 adults (ages 20-65) with a history of Long COVID

**Main measures:** The Composite Autonomic Symptom 31 (COMPASS-31) score, the RAND 36-Item Health Survey, the prevalence of new postural orthostatic tachycardia syndrome (POTS), and predictors of autonomic dysfunction, including POTS

**Key results:** 71.9% of Long COVID patients had a COMPASS-31 score >20, suggestive of moderate to severe autonomic dysfunction. The median symptom duration was 36 [30-40] months, reaching up to 3.5 years after SARS-CoV-2 infection. 37.5% of Long COVID patients could no longer work or had to drop out of school due to their Long COVID illness. 40.5% were newly diagnosed with POTS following SARS-CoV-2 infection.

**Conclusions:** Evidence of persistent moderate to severe autonomic dysfunction was seen in 71.9% of Long COVID patients in our study, with a 36-month median symptom duration, suggesting that enduring autonomic dysfunction is highly prevalent in the Long COVID population. Moderate to severe autonomic dysfunction was significantly correlated with impaired function and capacity, highlighting the need to address POTS and other manifestations of autonomic dysfunction as a key component of Long COVID management.

## Introduction

Autonomic dysfunction has been recognized as a common complication of Long COVID (LC), also known as post-acute sequalae of SARS-CoV-2 (PASC)^1^. However, despite the magnitude of the pandemic and the growing number of those with LC, longitudinal outcome data in those with autonomic dysfunction associated with LC are limited. Given the substantial level of disability in this population^2^, understanding the trajectory of autonomic symptoms is essential to better understand the natural history of the disease and inform prognosis. While several studies have reported longitudinal data^3,4,5^ in those with LC, studies focusing on autonomic symptom burden are limited. We previously reported on autonomic symptom severity in an adult LC cohort, findings symptoms of mild to moderate autonomic dysfunction in 66% of participants^1^. In this follow-up study, we recruited the same participants to complete follow-up surveys and orthostatic active stand testing to better assess LC symptom duration, severity, and impact on quality of life, as well as potential predictors for prolonged symptom duration.

## Materials and Methods

Participants were recruited from a prior study cohort, originally recruited through LC support groups and social media channels (October 2020 and August 2021), as previously described^1^. Dysautonomia support groups were excluded to avoid sampling bias towards severe autonomic dysfunction. Exclusion criteria included incomplete surveys, symptom duration < 90 days, symptom onset before November 2019, and age ≥65 years (due to concern of survivor bias secondary to disproportionately high mortality in this age range). Patients with pre-existing autonomic diagnoses were included in exploratory analyses but excluded when assessing LC-related autonomic dysfunction.

Data collection was conducted through the online Research Electronic Data Capture (REDCap) platform. All participants provided digital informed consent prior to initiating the survey. Surveys were completed between 7/06/23 and 08/15/23. The study received ethical approval from the Stanford University Institutional Review Board.

Participants were asked to complete many of the original survey instruments. These included the Composite Autonomic Symptom Score-31 (COMPASS-31) to assess symptoms of autonomic dysfunction, with scores ranging from 0-100 with greater scores indicating greater autonomic dysfunction. As in our baseline study,^1^ a score of ≥20 was established *a priori* as indicative of moderate-to-severe autonomic dysfunction. It is important to note, there are patients with a COMPASS-31 score less than 20 who experience clinically significant autonomic dysfunction. The Orthostatic Hypotension Questionnaire (OHQ) assesses symptoms of orthostatic intolerance, with higher scores ranging from 0-100 indicating greater symptom burden; the fatigue severity scale (FSS) assesses fatigue, with scores from 9-63, with ≥ 36 indicating abnormal fatigue; the General Anxiety Disorders assessment (GAD7) assesses anxiety symptoms, with scores ranging from 0 to 24 with cut-offs of 5, 10, and 15 representing mild, moderate, and severe levels of anxiety; and the RAND 36-item health survey (RAND-36) assesses quality of life through multiple health domains, each scored from 0-100 with lower scores indicating greater disability.

Non-standardized survey instruments were also administered to assess, among other variables, current symptoms, details of SARS-CoV-2 reinfection, vaccination status, past medical history, medications prescribed, and new post-COVID diagnoses. The ten most reported LC symptoms from our baseline survey^1^ were, in order of decreasing frequency: fatigue, brain fog, headache, shortness of breath with exertion, body aches, palpitations, lightheadedness, tachycardia, and difficulty sleeping. We, therefore, chose to focus on these ten symptoms in our follow-up questionnaire. Regarding past medical history, patients self-reported joint hypermobility by indicating whether their joints were “excessively flexible.” Participants were asked to list all the medications prescribed for their LC symptoms and identify the one that most effectively alleviated their symptoms. Participants reported whether they had received new diagnoses following their SARS-CoV-2 infection(s) and were specifically queried about diagnoses of mast cell activation syndrome (MCAS) and autonomic disorders, including postural tachycardia syndrome (POTS).

Finally, participants were instructed to complete a ten-minute active stand test with an automated sphygmomanometer cuff placed over the brachial artery. Participants measured blood pressure and heart rate in the supine position after a 10-minute supine rest period, followed by every minute standing for 10 minutes, or as long as tolerated. Exaggerated postural tachycardia was defined as an increase in heart rate (HR) of at least 30 bpm sustained over at least two consecutive measurements of standing in the absence of orthostatic hypotension (OH), as per consensus criteria^6^. Classic OH was defined as a blood pressure (BP) fall of at least 20 mmHg systolic or 10 mm Hg diastolic within the first 3 minutes of standing, sustained over at least 2 minutes, while delayed OH was defined as a similar sustained fall in BP occurring after 3 minutes of standing, as per consensus criteria^6^.

### Statistical Analysis

We employed a staggered approach in our analysis, estimating the adjusted associations between independent predictors and moderate-to-severe autonomic burden (COMPASS-31 score ≥ 20) using a multivariate logistic regression model (Supplementary Table 1). We also used a multivariable logistic regression to determine predictors of a POTS diagnosis following COVID infection. The analysis included eligible participants without prior dysautonomia diagnosis (N=489) (Supplementary Table 2).

Categorical variables are displayed as counts and percentages. Continuous variables, Gaussian and non-Gaussian, are shown as mean ± SD and median with interquartile range, respectively, as verified by the Shapiro-Wilk test. Gaussian variables’ comparisons used the t-test and Cohen’s d for effect size, while non-Gaussian variables used the Wilcoxon rank sum test. Categorical variables across groups were compared using χ2 or Fisher’s exact test (for counts less than 5 per category). Spearman correlation coefficients were calculated to evaluate the strength and relationship of associations between COMPASS-31 scores and other survey measures.

Correlation coefficients (r) exceeding ±0.5 were considered strong, and statistically significant associations were reported. We established a statistical threshold of p<0.05 in accordance with our prior study^1^ and applied false discovery rate analyses to correct for possible Type I errors. All statistical operations were conducted using Stata/BE 18.0^7^, Python 3.12.1^8^, and MATLAB R2023b^9^.

## Results

A total of 526 LC participants were included for final analysis, of which 86 participants provided orthostatic stand test results (Figure 1).

**Figure 1.**
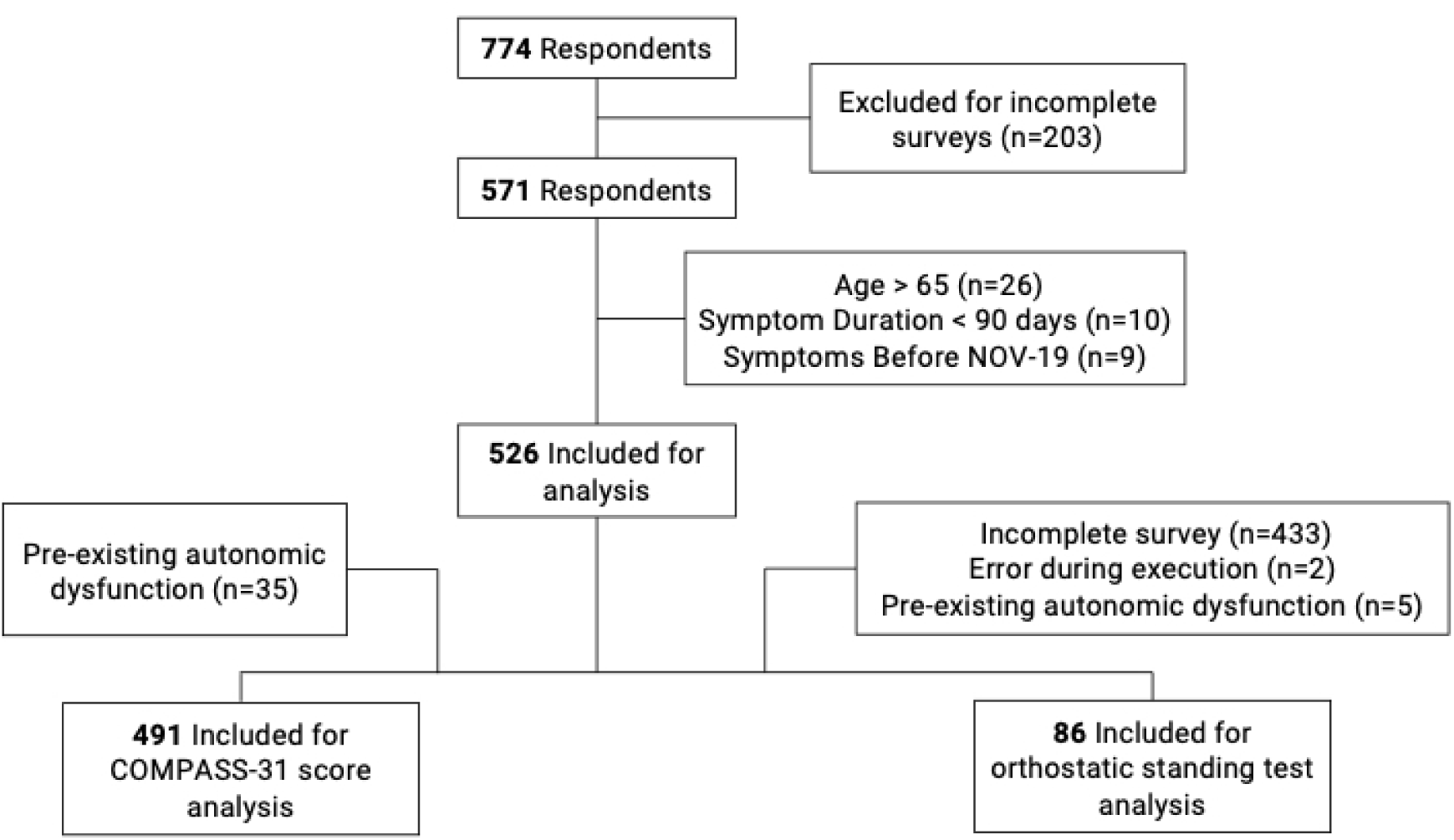
Participant flow diagram. 526 eligible surveys and 86 Active Standing test results were included in the study analysis.

### Demographics

Of the 526 participants, 467 (88.8%) reported female sex at birth (Table 1). The age of respondents ranged from 20-65 years old, with a median age of 48.5 [41-56] years. Eighty-four percent of participants identified as White, 6.7% as Biracial/Multi-Racial, 2.7% as Latino/Hispanic, 2.3% as Asian, and 1.5% as Black. Participants identifying as Native American/Indigenous, Middle Eastern, or Pacific Islander each comprised less than 1% of the cohort. Most survey participants resided in the USA (79.7%), with the greatest number from California (Figure 2).

**Figure 2.**
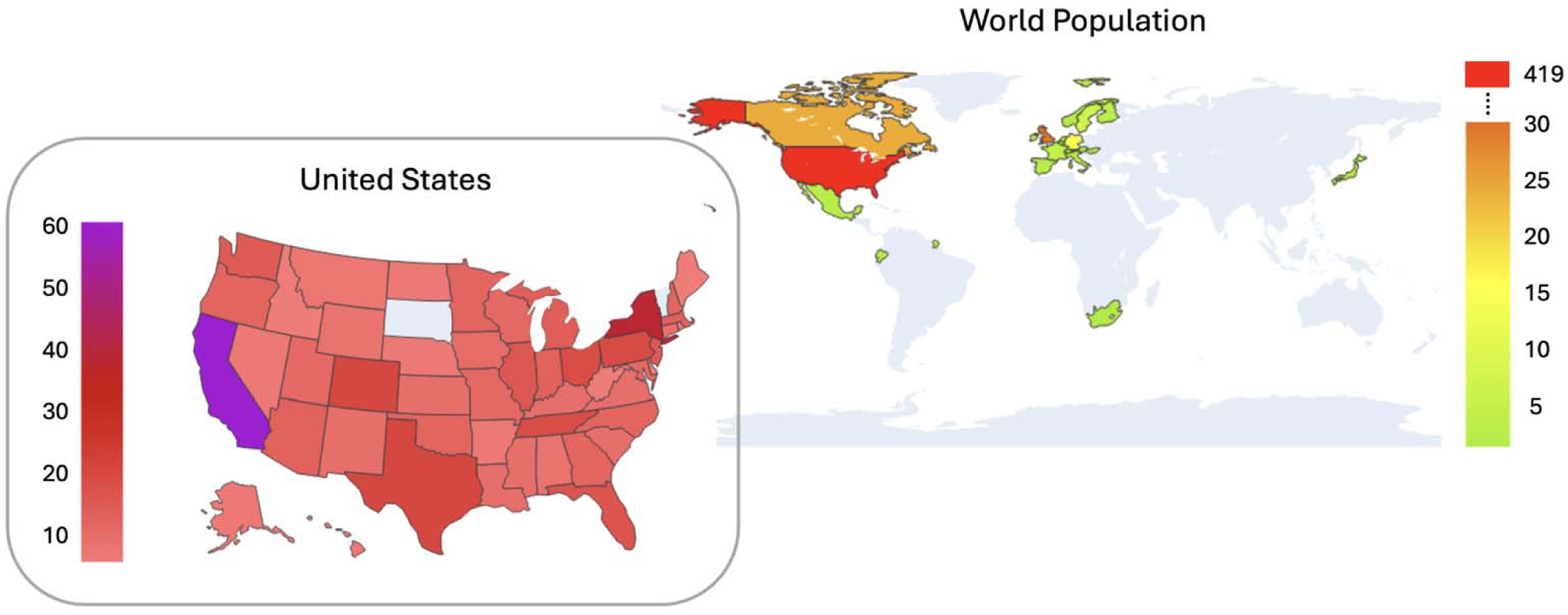
Geographical distribution of study participants reporting residence in the U.S. (top) and a country outside of the U.S. (bottom) (count) (N=363).

**Table 1.** Participant demographics and past medical history.

| Characteristic | N=526 |
| --- | --- |
| **Assigned Sex at Birth** |  |
| Female | 467 (88.8) |
| Male | 59 (11.2) |
| **Age** |  |
| 20-35 | 57 (10.8) |
| 36-45 | 151 (28.7) |
| 46-55 | 185 (35.2) |
| 56+ | 133 (25.3) |
| Mean (SD) | 48.1 (9.9) |
| Median [IQR] | 48.5 [41-56] |
| **Race** |  |
| White | 441 (83.8) |
| Biracial/Multiracial | 35 (6.65) |
| Latino/Hispanic | 14 (2.66) |
| Asian | 12 (2.28) |
| Black | 8 (1.52) |
| Native American/Indigenous | 3 (0.57) |
| Middle Eastern | 4 (0.76) |
| Pacific Islander | 1 (0.19) |
| *Not Disclosed* | 8 (1.52) |
| **Country** |  |
| US | 419 (79.7) |
| Outside US | 106 (20.2) |
| *Not Disclosed* | 1 (0.1) |
| **Past Medical History** |  |
| *Prior Autoimmune Dx* |  |
| Yes | 80 (15.2) |
| No | 446 (84.8) |
| *Prior Autonomic Dx* |  |
| Yes | 35 (6.7) |
| No | 491 (93.4) |
| **Hypermobility<sup>a</sup>** |  |
| Yes | 156 (29.7) |
| No | 370 (70.3) |

| COVID-19 History |  |
| --- | --- |
| **COVID-19 Testing Method**<br>Confirmed Medical PCR<br>At Home Rapid Antigen Test<br>Dx by Antibodies<br>Dx by Symptoms only | 293 (55.7)<br>15 (2.9)<br>37 (7.0)<br>181 (34.4) |
| **Vaccinated at time of COVID infection<sup>2</sup>**<br>Yes<br>No | 512 (97.3%)<br>24 (2.7%) |
| PASC Symptom Prevalence and Duration |  |
| **Current PASC Symptoms<sup>3</sup>**<br>Yes<br>No<br>Not Sure | 450 (85.6)<br>31 (5.89)<br>45 (8.56) |
| **PASC Symptom Duration** | Mean= 32.7 (SD= 10.0)<br>Median= 36 [range: 3-43] |
| Survey Questions:<br><sup>1</sup> Are your joints excessively flexible (hypermobile)?<br><sup>2</sup> Were you fully vaccinated when you were diagnosed with the COVID infection that led to Long-COVID?<br><sup>3</sup> Do you still experience symptoms that you or your doctor attribute to a long COVID illness? |  |

### Past Medical History

Eighty (15.2%) participants reported a history of an autoimmune diagnosis prior to their first SARS-CoV-2 infection (most common diagnoses including Hashimoto’s thyroiditis, celiac disease, Sjogren’s disease, rheumatoid arthritis, systemic lupus erythematosus and psoriasis). Thirty-five (6.7%) participants reported an autonomic diagnosis that preceded their first SARS-CoV-2 infection. Of that group, 25 (4.8%) reported a prior POTS diagnosis. One hundred and fifty-six (29.7%) patients reported joint hypermobility (Table 1).

### COVID-19 & Vaccination History

Participants were categorized into four groups based on their reported COVID-19 diagnosis method (Table 1). Importantly, this cohort includes patients diagnosed with COVID-19 early in the pandemic, where testing was not as widely available as it is today. Five hundred and twelve (97.3%) patients were unvaccinated at the time of their incident SARS-Co-V 2 infection that led to their LC illness. At the time of follow-up survey completion, 424 (80.6%) participants reported being fully vaccinated, 60 (11.4%) reported being partially vaccinated, and 42 (8.0%) were unvaccinated (Table 1).

### LC Symptoms and Duration

Four hundred and fifty (85.6%) participants reported that, at the time of follow-up survey, they were still experiencing symptoms that they and/or their doctor attributed to LC, with a median symptom duration of 36 months [30-40] after the onset of their infection (Table 1). The most frequently reported persistent LC symptoms, were fatigue (88.8%), brain fog (81.4%), and difficulty sleeping (70.5%) (Table 2). Lightheadedness was reported by 64.8%, palpitations by 58.9% and tachycardia by 54.4% of participants.

**Table 2.** Symptom prevalence.

| Symptom | Count (%) |
| --- | --- |
| Fatigue | 467 (88.8) |
| Brain Fog | 428 (81.4) |
| Difficulty Sleeping | 371 (70.5) |
| Headache | 363 (69.0) |
| Body Aches | 357 (67.9) |
| Shortness of Breath with Exertion | 342 (65.2) |
| Lightheadedness | 341 (64.8) |
| Palpitations | 310 (58.9) |
| Tachycardia | 286 (54.4) |
| Other* | 195 (37.1) |
| None | 18 (3.4) |
| * Most commonly reported "other" symptoms include: post-exertional malaise (PEM), joint pain, GI symptoms, nerve pain, paresthesias, tinnitus |  |

### Effect of SARS-CoV-2 Reinfection on Long COVID Symptom Severity

Of the 309 (58.8%) participants who reported at least one SARS-CoV-2 reinfection, 138 (44.7%) experienced worsening of LC symptoms, 116 (37.5%) reported no change, and 55 (17.8%) saw an improvement. A Pearson Chi-squared test was conducted to evaluate the relationship between symptom reinfection and the COMPASS-31 threshold score. The results indicated a statistically significant association (χ^2^ = 15.63, p=0.016), suggesting that reinfection influences the severity of LC autonomic dysfunction.

### Effect of COVID-19 Vaccination on Long COVID Symptom Severity

Most participants (97.34%) were not vaccinated at the time of their first SARS-CoV-2 infection, while 14 (2.66%) participants had at least one COVID-19 vaccination before their first SARS-CoV-2 infection. Four hundred and eighty-four participants (92.0%) reported receiving at least one COVID-19 vaccination at the time of the follow-up survey. Following COVID-19 vaccination, 60.54% reported no change in LC symptoms, while 20.25% reported improvement, and 19.21% reported worsening of LC symptoms.

### Autonomic, Fatigue, and Anxiety Scores

Of the 491 Long COVID patients with no prior history of an autonomic disorder, 353 (71.9%) had a COMPASS-31 score ≥ 20, suggestive of moderate to severe autonomic dysfunction (median 32.3 [18.0-44.4]). Thirty-five participants were excluded from this analysis due to pre-existing autonomic disorders.

Comparing demographics and past medical history of those with COMPASS-31 scores ≥ 20 to those with scores < 20 revealed several significant differences (Table 3), with female sex (p=0.012) and joint hypermobility (p=0.019) imparting greater risk of more severe autonomic symptoms following COVID-19 infection.

**Table 3.** Characteristics and relevant medical history of participants based on COMPASS-31 score. (N=491)

| Characteristic | COMPASS-31<br>< 20<br>N = 138 (27.38) | COMPASS-31<br>≥ 20<br>N = 353 (72.62) | P-value<br>(Chi Square/ Fisher's Exact<br>Test/ Wilcoxon Ranksum) |
| --- | --- | --- | --- |
| **Age**<br>20-35<br>36-45<br>46-55<br>56+ | 9 (6.5)<br>32 (23.2)<br>63 (45.7)<br>35 (24.6) | 39 (11.1)<br>108 (30.6)<br>113 (32.0)<br>98 (26.4) | 0.026* |
| Mean (SD)<br>Median [IQR] | 49.1 (9.24)<br>50 [42-55] | 47.7 (10.1)<br>48 [41-56] | 0.165** |
| **Assigned Sex at Birth**<br>Female<br>Male | 114 (82.6)<br>24 (17.4) | 320 (90.65)<br>33 (9.4) | 0.012* |
| **Race**<br>White<br>Black | 115 (83.3) | 300 (85.0) | 0.941* |
| Hispanic/Latino | 3 (2.2) | 5 (1.4) |  |
| Asian | 4 (2.9) | 7 (2.0) |  |
| Native American/Indigenous | 4 (2.9) | 8 (2.3) |  |
| Middle Eastern | 0 (0) | 3 (0.9) |  |
| Pacific Islander | 1 (0.7) | 2 (0.6) |  |
| Biracial/Multiracial | 0 (0) | 1 (0.3) |  |
| *Not Disclosed* | 8 (5.8) | 22 (6.2) |  |
|  | 3 (2.2) | 5 (1.4) |  |
| **Ethnicity** |  |  |  |
| Hispanic | 7 (5.1) | 19 (5.3) | 0.892* |
| Non-Hispanic | 128 (92.7) | 328 (92.9) |  |
| *Not Disclosed* | 3 (2.2) | 6 (1.7) |  |
| **Country** |  |  |  |
| US | 81 (58.7) | 257 (72.8) | 0.002* |
| Outside US | 57 (41.3) | 96 (27.2) |  |
| **Medical History** |  |  |  |
| **Any Autoimmune Dx** |  |  |  |
| Yes | 14 (10.1) | 52 (14.7) | 0.181* |
| No | 124 (89.9) | 301 (85.3) |  |
| **Hypermobility<sup>†</sup>** |  |  |  |
| Yes | 27 (19.6) | 106 (30.0) | 0.019* |
| No | 111 (80.4) | 247 (70.0) |  |
| **COVID-19 History** |  |  |  |
| **Vaccinated at time of COVID Infection** |  |  |  |
| Yes | 4 (2.9) | 7 (2.0) | 0.54* |
| No | 134 (97.1) | 346 (98.0) |  |
| **Positive COVID-Testing Method** |  |  |  |
| Confirmed Medical PCR | 71 (51.4) | 204 (57.8) |  |
| At Home PCR | 3 (2.2) | 10 (2.8) |  |
| Dx by Antibodies | 11 (8.0) | 23 (6.5) |  |
| Dx by Symptoms only | 53 (38.4) | 116 (32.9) |  |
| **Vaccinated<sup>‡</sup>** |  |  |  |
| Yes | 131 (94.9) | 323 (91.5) | 0.196* |
| No | 7 (5.1) | 30 (8.5) |  |
| **Booster** |  |  |  |
| Yes (monovalent) | *N*=131<br>44 (33.6) | *N*=323<br>116 (35.9) | 0.074* |
| Both (mono & bi-valent) | 77 (58.8) | 160 (49.5) |  |
| No | 10 (7.6) | 47 (14.55) |  |

| Vaccine Type |  |  |  |
| --- | --- | --- | --- |
| Pfizer | 67 (48.6) | 184 (52.1) | 0.002* |
| Moderna | 48 (36.4) | 105 (29.8) |  |
| J&J | 3 (2.2) | 23 (6.5) |  |
| AstraZeneca | 13 (9.4) | 8 (2.3) |  |
| Other | 0 | 3 (0.85) |  |
| None | 7 (5.1) | 30 (8.5) |  |
| <sup>1</sup> Are your joints excessively flexible (hypermobile)? |  |  |  |
| <sup>2</sup> Have you had a COVID Vaccine? |  |  |  |
| *A chi-square test |  |  |  |
| ** Wilcoxon Rank Sum |  |  |  |

Multivariable logistic regression of the 491 participants with no prior history of an autonomic disorder (Supplementary Table 1) also indicated that female sex and joint hypermobility were significant predictors of more severe autonomic dysfunction, defined by a COMPASS-31 total score ≥ 20. Those who reported female sex had 227% increased odds of moderate to severe autonomic dysfunction compared to those who reported male sex (OR: 0.44, p=0.009), while those with joint hypermobility had 68.0% increased odds of moderate to severe autonomic dysfunction (OR: 1.68, p=0.044) compared to those without joint hypermobility. Prior autoimmune diagnoses, ethnicity, and vaccination status at the time of infection were not significant predictors of autonomic dysfunction. However, it should be noted that the sample size of non-white participants was limited, and most participants were unvaccinated at the time of their initial infection.

The follow-up cohort’s median OHQ score was 46.0 [26.0, 65.0], suggesting a moderate burden of orthostatic intolerance. Participants with COMPASS-31 scores ≥20 had higher OHQ scores compared to those with scores <20 (56.0 [37.0, 71.0] vs. 25.5 [9.75, 37.0], p<0.0001), consistent with prior findings^1^ and suggesting that those with higher COMPASS-31 scores were driven at least in part by orthostatic intolerance (Figure 3).

**Figure 3.**
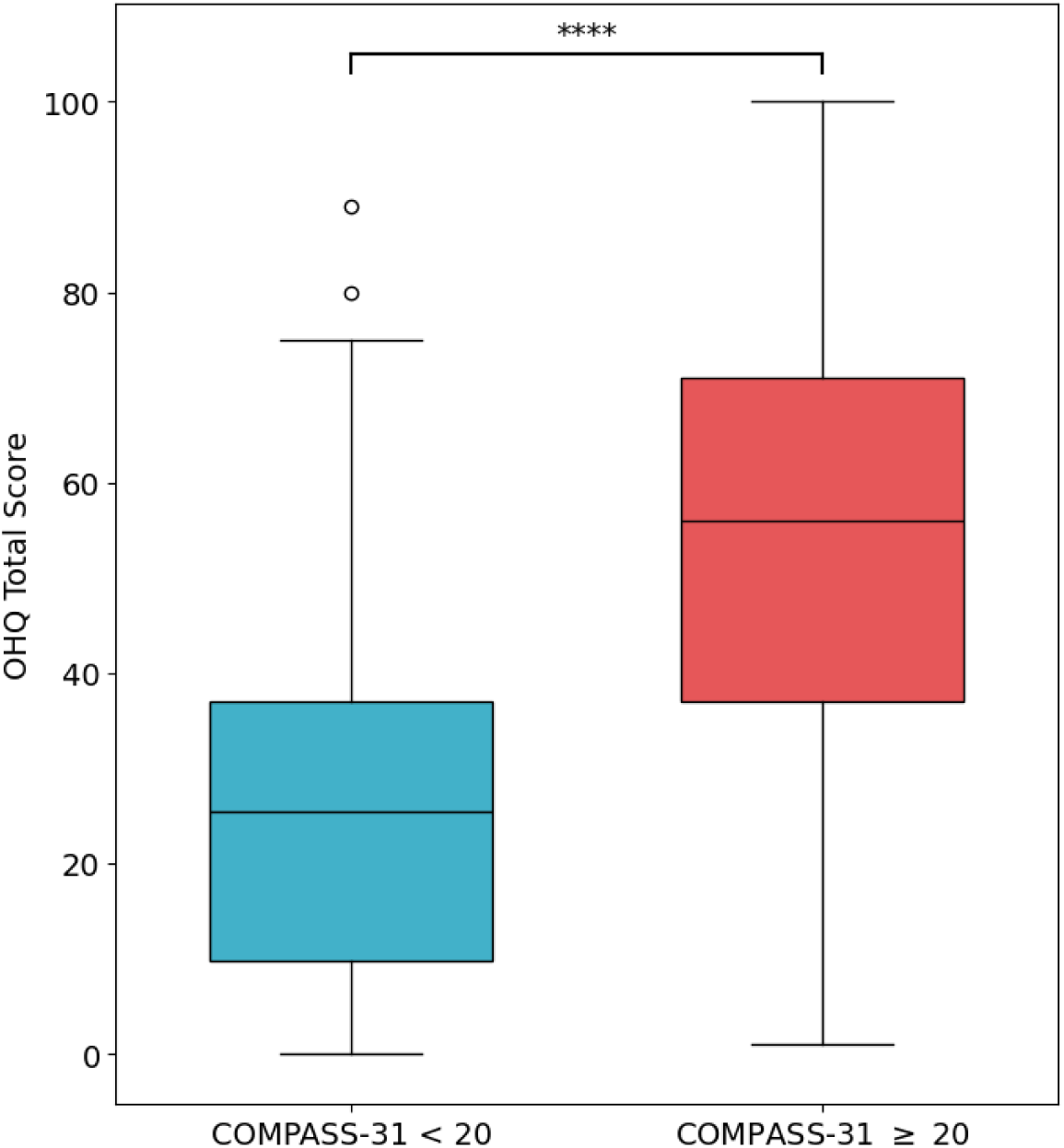
OHQ Scores by COMPASS-31 Score group.

The median FSS score was 6.0 [4.56, 6.67]. Our cohort’s mean FSS score (5.42 ± 1.61) was higher than in healthy individuals (2.6 ± 1.1)^10^. Participants with COMPASS-31 scores ≥20 had higher FSS scores than those with scores <20 (6.33 [5.22, 6.78] vs. 4.7 [3.22, 6.0], p<0.0001). A moderate positive correlation (ρ = 0.315) existed between COMPASS-31 orthostatic subdomain scores and FSS scores.

The average GAD-7 score was 6.58 ± 5.32, with 41.83% reporting minimal, 34.22% mild, 13.9% moderate, and 10.1% severe anxiety. Participants with COMPASS-31 scores ≥20 had higher GAD-7 scores [6.0 (3.0, 10.0)] than those with scores <20 [4.0 (1.0, 7.0), p<0.0001].

### Impact of Autonomic Symptom Burden on Disability

Of all study participants, 197 (37.5%) could no longer work or had to drop out of school due to their illness. Among those with a COMPASS-31 ≥ 20 (n=353), 152 (43.1%) could no longer work or had to drop out of school due to their illness, which was significantly higher than the 26.1% observed in those with scores < 20 (p=0.001). (Table 4)

**Table 4.**
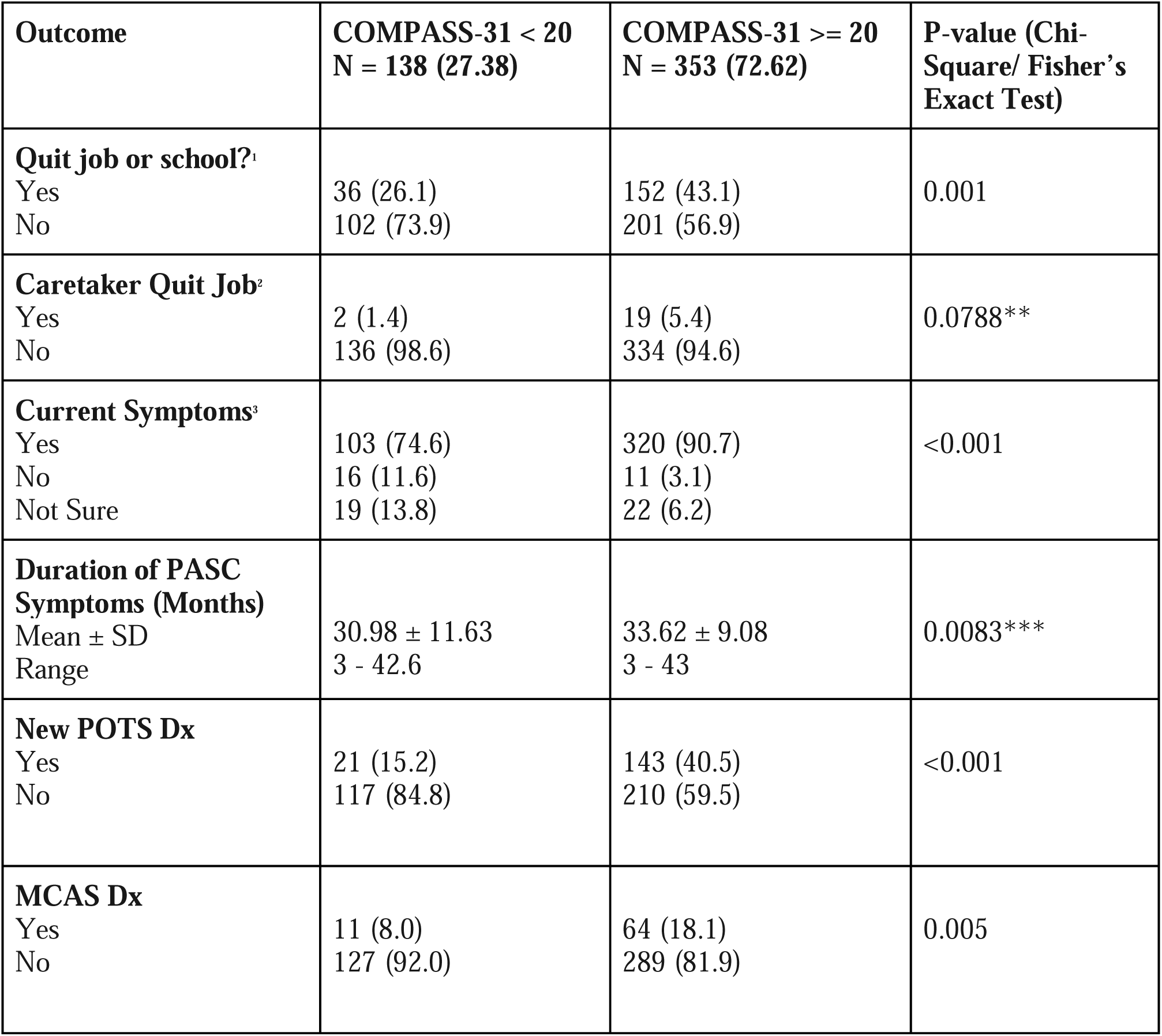
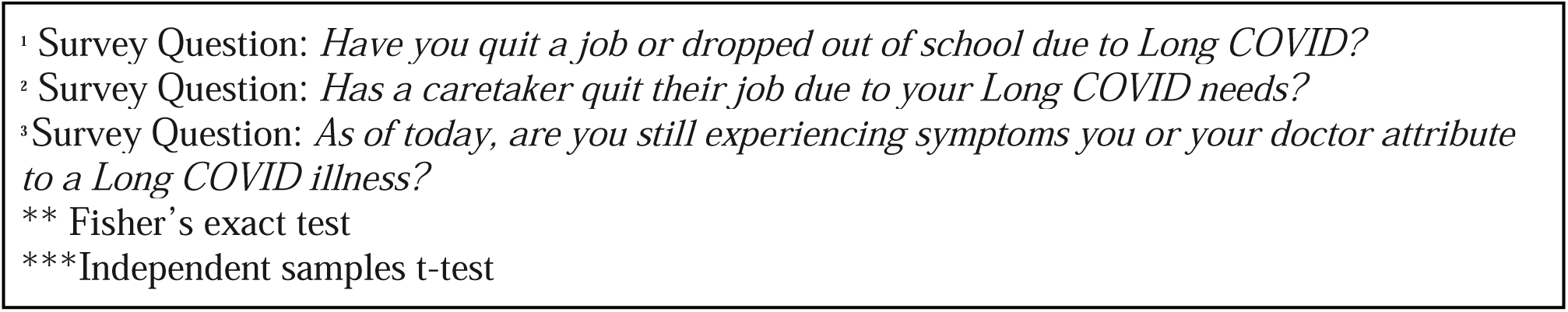
Health and diagnostic outcomes based on COMPASS-31 categorization.

| **Outcome** | **COMPASS-31 < 20**<br>**N = 138 (27.38)** | **COMPASS-31 >= 20**<br>**N = 353 (72.62)** | **P-value (Chi-Square/ Fisher's Exact Test)** |
| --- | --- | --- | --- |
| **Quit job or school?<sup>1</sup>** |  |  |  |
| Yes | 36 (26.1) | 152 (43.1) | 0.001 |
| No | 102 (73.9) | 201 (56.9) |  |
| **Caretaker Quit Job<sup>2</sup>** |  |  |  |
| Yes | 2 (1.4) | 19 (5.4) | 0.0788** |
| No | 136 (98.6) | 334 (94.6) |  |
| **Current Symptoms<sup>3</sup>** |  |  |  |
| Yes | 103 (74.6) | 320 (90.7) | <0.001 |
| No | 16 (11.6) | 11 (3.1) |  |
| Not Sure | 19 (13.8) | 22 (6.2) |  |
| **Duration of PASC Symptoms (Months)** |  |  |  |
| Mean ± SD | 30.98 ± 11.63 | 33.62 ± 9.08 | 0.0083*** |
| Range | 3 - 42.6 | 3 - 43 |  |
| **New POTS Dx** |  |  |  |
| Yes | 21 (15.2) | 143 (40.5) | <0.001 |
| No | 117 (84.8) | 210 (59.5) |  |
| **MCAS Dx** |  |  |  |
| Yes | 11 (8.0) | 64 (18.1) | 0.005 |
| No | 127 (92.0) | 289 (81.9) |  |

<sup>1</sup> Survey Question: *Have you quit a job or dropped out of school due to Long COVID?* <sup>2</sup> Survey Question: *Has a caretaker quit their job due to your Long COVID needs?* <sup>3</sup> Survey Question: *As of today, are you still experiencing symptoms you or your doctor attribute to a Long COVID illness?* \*\* Fisher's exact test \*\*\*Independent samples t-test

Our cohort demonstrated markedly lower scores across *all* RAND-36 health domains compared to established population means^11^ (Supplementary Table 3), reflecting increased disability associated with LC. When analyzing by COMPASS score, those with a COMPASS-31 score ≥20 reported significantly worse outcomes on almost all scales (Supplementary Table 4).

A significant negative correlation was observed between COMPASS-31 scores and all RAND- 36 health domains (Figure 4), with the strongest associations being physical functioning and pain, where greater autonomic dysfunction was associated with higher disability. Fatigue and general health also showed inverse relationships with COMPASS-31, further emphasizing that greater autonomic dysfunction contributes to lower quality of life in LC patients.

**Figure 4.**
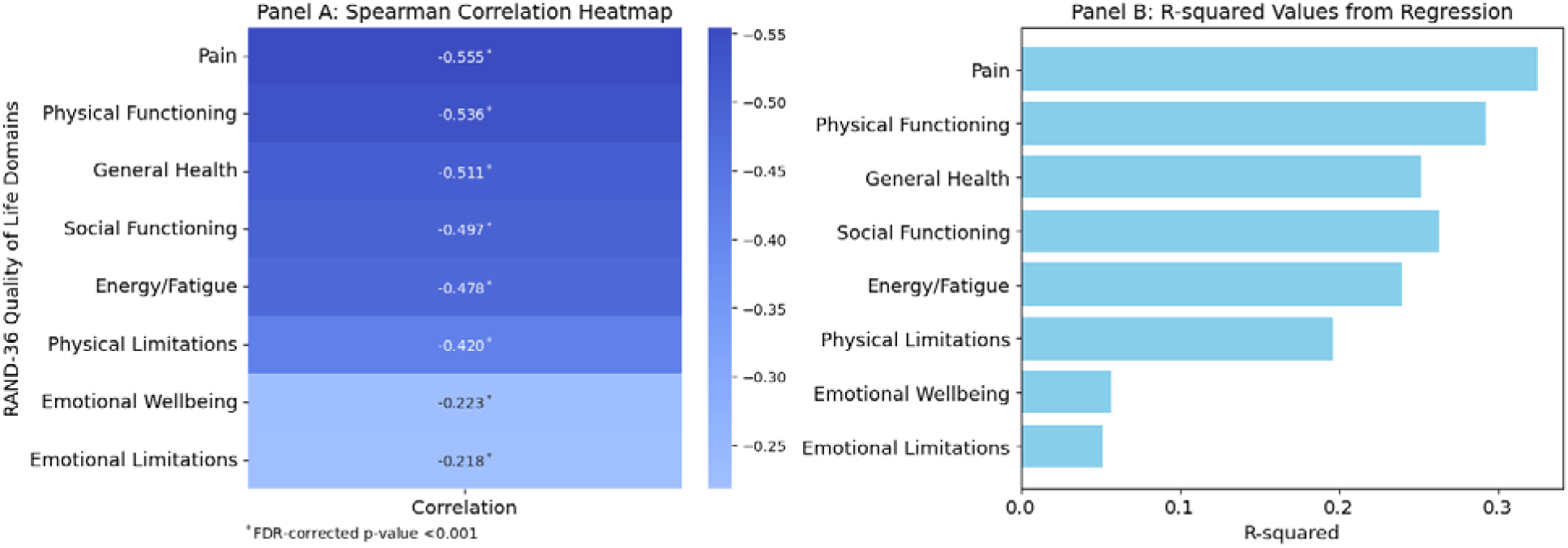
Relationship between COMPASS-31 scores and RAND-36 Quality of Life Domains

### New Autonomic Diagnoses and Risk Factors for POTS

Of the 491 patients with no prior history of an autonomic disorder, 214 (43.58%) reported a new autonomic disorder diagnosis following their first SARS-CoV-2 infection. Within this group, 161 (79.70%) reported a new diagnosis of POTS, while 51 (25.25%) reported a new diagnosis of inappropriate sinus tachycardia (IST), and 22 (10.89%) reported a new diagnosis of orthostatic hypotension (OH). Among participants without a pre-existing autonomic disorder, 35 (6.5%) reported a new autonomic diagnosis following SARS-CoV-2 reinfection, with 24 (68.57%) of these participants reporting a new diagnosis of POTS. Of all participants who reported new autonomic diagnoses post-COVID, only 32 (13.6%) reported that their autonomic disorder had resolved at the time of follow-up survey.

Eighty-three (15.8%) of participants reported a diagnosis of MCAS following SARS-CoV-2 infection. Those with COMPASS-31 scores ≥ 20 were significantly more likely to have received an MCAS diagnosis (18.1%) compared to those with lower COMPASS-31 scores (8.0%) (p=0.005).

Multivariable logistic regression identified several risk factors for developing POTS in our cohort. The most robust risk factor identified, was joint hypermobility, which imparted 200% increased odds of developing POTS following after COVID-19 (OR = 2.0502, 95% CI = 1.3390-3.1389, p = 0.001). In addition, those ≥ 55 years of age had 49% decreased odds of developing POTS compared to those ≤ 40 years of age (OR = 0.51, 95% CI = 0.292-0.891, p =0.018). Other variables controlled for in the regression include sex, ethnicity, vaccination status at the time of SARS-CoV-2 infection, and country of origin, which were not predictive of LC-POTS diagnoses.

### Medications

Of all therapies queried, participants indicated beta-blockers (15.9%) as the most effective for reducing LC-related symptoms (Table 5). Other therapies perceived as beneficial included low-dose naltrexone (LDN) (4.18%), steroids (1.9%, ivabradine (3.2%), wake-promoting agents or stimulants (2.7%), and intravenous saline (1.9%). Most participants found that no single medication was most effective at reducing their symptoms. (Table 5)

**Table 5.** Patient-reported medications prescribed for LC symptoms.

| Medication/Treatment | Ever Prescribed* | Most Helpful** |
| --- | --- | --- |
| None of these | 157 (29.9) | 248 (47.2) |
| Beta-Blockers | 194 (36.9) | 82 (15.9) |
| Low Dose Naltrexone | 102 (19.4) | 22 (4.18) |
| Steroids | 145 (27.6) | 20 (3.4) |
| Ivabradine | 46 (8.8) | 17 (3.2) |
| Wake-Promoting Agents/Stimulants | 64 (12.2) | 14 (2.7) |
| IV Saline | 59 (11.2) | 10 (1.9) |
| Midodrine | 47 (8.9) | 9 (1.7) |
| Pyridostigmine | 37 (7.0) | 7 (1.3) |
| IVIG | 22 (4.2) | 7 (1.3) |
| Fludrocortisone | 36 (6.84) | 3 (0.6) |
| Droxidopa | 4 (0.8) | 2 (0.4) |
| Central Alpha Blockers (Clonidine, Guanfacine) | 24 (4.6) | 1 (0.2) |
| *Other Not Listed* | *192 (36.5)* | *84 (16.0)* |
| *Values exceed 100% as participants could report multiple prescribed medications.<br>**Survey Questions: Which has been the single most helpful medication in treating your symptoms? |  |  |

**Table 6.**
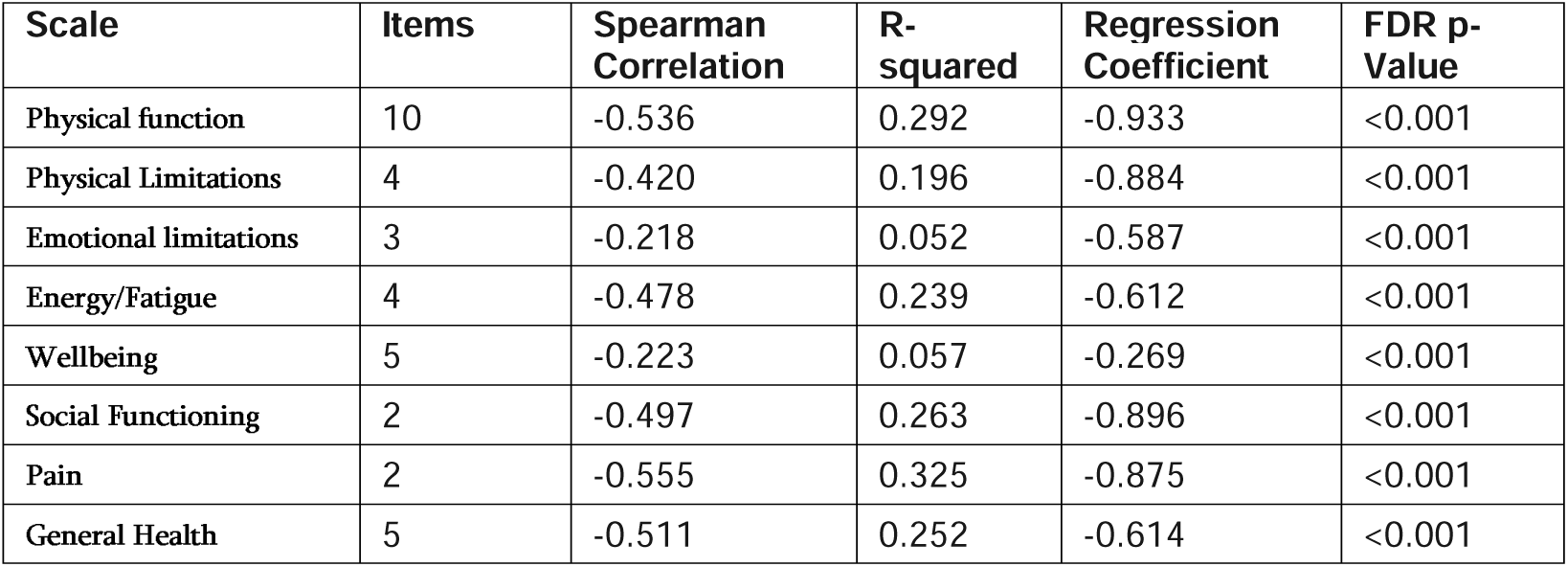
RAND-36 domains correlated to total COMPASS-31 scores. Correlation analysis of health-related quality of life measures compared to COMPASS-31 scores from survey participants: correlation and regression with statistical significance.

| Scale | Items | Spearman Correlation | R-squared | Regression Coefficient | FDR p-Value |
| --- | --- | --- | --- | --- | --- |
| Physical function | 10 | -0.536 | 0.292 | -0.933 | <0.001 |
| Physical Limitations | 4 | -0.420 | 0.196 | -0.884 | <0.001 |
| Emotional limitations | 3 | -0.218 | 0.052 | -0.587 | <0.001 |
| Energy/Fatigue | 4 | -0.478 | 0.239 | -0.612 | <0.001 |
| Wellbeing | 5 | -0.223 | 0.057 | -0.269 | <0.001 |
| Social Functioning | 2 | -0.497 | 0.263 | -0.896 | <0.001 |
| Pain | 2 | -0.555 | 0.325 | -0.875 | <0.001 |
| General Health | 5 | -0.511 | 0.252 | -0.614 | <0.001 |

### Active Stand Test Results

Of the 526 individuals who completed all follow-up surveys, 93 (17.68%) completed the active stand test. Seven participants were excluded from the analysis due to pre-existing autonomic diagnoses and incomplete surveys, leaving 86 participants (16.35%) with active stand test results for analysis (Supplementary Table 5).

34 (37.8%) participants displayed evidence of OH (12 with classic OH, and 22 with delayed OH). These participants were excluded from the subsequent analysis applying the POTS HR criteria. 33 (38.4%) participants met the criteria for an exaggerated postural tachycardia upon standing. The occurrence of exaggerated postural tachycardia was more prevalent in the COMPASS-31 ≥ 20 group, with 29 (55.77%) individuals meeting this criterion, compared to 4 (7.69%) participants in the COMPASS-31 < 20 group.

As expected, the median increase in HR between supine and standing was significantly greater in those who reported a new POTS diagnosis when compared to those who did not (40 [35.75, 52.5] vs.16 [11, 20], p<0.001). Of the 33 patients who completed the active stand test and met the criteria for an exaggerated postural tachycardia, 22 (67.67%) reported that they received a POTS diagnosis following a COVID-19 infection. 5 (15.15%) participants reported a diagnosis of IST and 2 (6.06%) participants were diagnosed with neurally mediated syncope.

## Discussion

Evidence of moderate to severe autonomic dysfunction was seen in 71.9% of LC participants in our cohort, at a median symptom duration of 36 months after infection, suggesting that chronic autonomic dysfunction is common in LC. We found that the severity of autonomic dysfunction, as defined by COMPASS-31 total scores, was directly associated with functional disability, with a significantly greater proportion of participants unable to work or complete their studies due to LC. These findings contrast with our prior baseline study, in which functional disability was not associated with COMPASS-31 scores^1^. One possible explanation is the contribution of reverse survivorship bias, whereby those in the baseline cohort whose LC symptoms improved or resolved were lost to follow-up (approximately 75% of participants failed to respond to follow-up survey requests), possibly enriching our follow-up cohort with more severely affected participants. Another possibility is the longer duration of follow-up, during which ongoing LC symptoms may have led to cumulative disability.

Factors associated with a greater risk of autonomic dysfunction included female sex and joint hypermobility. Our multivariate logistic regression analysis indicated that those with joint hypermobility had a 68.0% increased risk of moderate to severe autonomic dysfunction (OR: 1.68, p=0.044), while female participants had a 227% increased risk compared to males (OR: 0.44, p=0.009). Joint hypermobility was also associated with a 200% increased risk of a new POTS diagnosis. This is consistent with prior research finding that 24% of POTS patients have generalized joint hypermobility and 31% have hypermobile Ehlers Danlos Syndrome(hEDS)^12^. Connective tissue laxity may increase risk of venous pooling and reduced venous return, thus leading to postural tachycardia, however this has not been consistently demonstrated in hEDS pathophysiological studies. Although participants were not explicitly queried about EDS diagnoses, several participants volunteered this information, highlighting the need for further research on the interplay of joint hypermobility, female sex, and autonomic dysfunction in infection-associated chronic conditions like LC.

Forty-four percent of our cohort reported new autonomic diagnoses following their first SARS-CoV-2 infection, with POTS being the most common diagnosis, followed by IST and OH. Risk factors for new diagnoses included younger age, female sex, and joint hypermobility. Future clinical trials should consider stratifying those with generalized joint hypermobility or diagnoses of connective tissue disorders such as EDS.

Of the participants who completed active stand testing, most had COMPASS-31 scores ≥ 20. As expected, significant differences were observed in heart rate responses between those diagnosed with POTS and those without (40 [35.75, 52.5] vs. 16 [11, 20], p<0.001).

Among those without a history of a prior autonomic disorder, 33% reported a new POTS diagnosis, and within that cohort, 38% of those with active stand test results had an exaggerated postural tachycardia. Other studies have estimated a broad range of POTS rates in LC, between 15-80%^13–15^. All of these studies are confounded in some way or another, usually by referral bias, in which those LC patients with symptoms of POTS are referred for evaluation and testing.

However, one study that performed active stand testing in a cohort of 70 LC patients regardless of symptoms, finding that 30% had an exaggerated postural tachycardia^13^, findings very similar to our results. While true prevalence rates of POTS in LC remain to be determined, taken together, these findings suggest that up to one in three patients with LC are at risk of developing POTS, especially female patients with joint hypermobility.

The effect of SARS-CoV-2 reinfection on LC symptom severity was notable, with 44.7% of participants reporting worsened symptoms after reinfection. Conversely, vaccination had varied effects, with 60.54% reporting no change, 20.25% reporting improvement, and 19.21% reporting worsening of LC symptoms. These findings suggest that reinfection may exacerbate symptoms, whereas the impact of vaccination is more heterogeneous^16^ and warrants further investigation.

Despite the significant burden of disability in our cohort, few medications were reported to be effective at controlling autonomic symptoms. Beta-blockers were reported as the most effective medication, for 15.9% of participants. Most participants indicated that no single medication was highly effective in managing their symptoms. This underscores the need for more effective therapies and clinical trials in LC, an urgent unmet need that requires more than repurposing existing therapies.

## Limitations

Our findings may be influenced by survivor bias, as 65.6% of the initial sample didn’t respond to follow-up. Selection bias may also exist, with those more severely affected being more likely to participate. Survey limitations and recall bias could also affect self-reported data. Regarding the active stand test, patients were not instructed to discontinue medications and were not supervised by professionals, thus leading to an underestimate of rates of postural tachycardia in our cohort. However, we feel these limitations counterbalanced by the value of a longitudinal survey administered to a global population of LC patients, as well as the opportunity to include those infected early in the pandemic.

## Conclusions

Evidence of persistent moderate to severe autonomic dysfunction was seen in 71.9% of LC patients in our study, a median of 36 months after initial infection, suggesting that chronic autonomic dysfunction is highly prevalent in the LC population. Moderate to severe autonomic dysfunction was significantly correlated with impaired function and capacity, highlighting the need to address autonomic dysfunction as a key component of LC management. POTS was the most common autonomic diagnosis reported, affecting approximately one in three LC participants. This finding was supported by similar rates of exaggerated postural postural tachycardia on active stand testing.

## Data Availability

All data produced in the present study are available upon reasonable request to the authors.

## Funding

This research did not receive any specific grant from funding agencies in the public, commercial, or not-for-profit sectors.

## Glossary

*Glossary -*
| Term | Definition |
| --- | --- |
| Autonomic Dysfunction | A disorder where the autonomic nervous system malfunctions, affecting involuntary functions like heart rate, blood pressure, and sweating. |
| Beta-Blockers | A class of medications commonly used to manage cardiovascular conditions by blocking the effects of adrenaline on the heart (reduction in blood pressure and heart rate). |
| Composite Autonomic Symptom Score-31 (COMPASS-31) | A 31-item questionnaire assessing the severity of autonomic symptoms across multiple systems, with scores from 0 to 100. |
| COVID-19 (Coronavirus Disease 2019) | A viral respiratory illness caused by the SARS-CoV-2 virus. |
| Ehlers-Danlos Syndrome (EDS) | A group of genetic disorders affecting connective tissues, often resulting in joint hypermobility, skin elasticity, and tissue fragility. |
| Exaggerated Postural Tachycardia | An abnormal, excessive increase in heart rate that occurs upon standing. |
| Fatigue Severity Scale (FSS) | A 9-item questionnaire measuring the impact of fatigue on daily activities, with scores ranging from 9 to 63. |
| General Anxiety Disorder-7 (GAD7) | A 7-item questionnaire for screening and measuring the severity of generalized anxiety disorder, with scores ranging from 0 to 21. |
| Inappropriate Sinus Tachycardia (IST) | A condition where the heart rate is |
|  | persistently elevated or increases disproportionately with minor exertion. |
| Joint Hypermobility | A medical condition where joints easily move beyond the normal range of motion expected for a particular joint. |
| Logistic Regression | A statistical analysis used to model the relationship between a binary, dependent variable and one or more independent predictor variables. |
| Long-COVID (LC) | A clinically diagnosed condition characterized by persistent symptoms and long-term health issues following a COVID-19 infection; also known as post-acute sequelae of SARS-CoV-2 (PASC). |
| Low-Dose Naltrexone (LDN) | A medication used to modulate the immune system and potentially treat chronic pain and autoimmune conditions. |
| Mast Cell Activation Syndrome (MCAS) | A condition where mast cells inappropriately release chemical mediators, causing multisystem symptoms affecting organ systems such as gastrointestinal, cardiovascular, and respiratory. |
| Multivariate Logistic Regression | An extension of logistic regression that considers multiple independent variables simultaneously to predict the probability of a binary outcome. |
| Orthostatic Hypotension Questionnaire (OHQ) | A questionnaire to evaluate the symptoms and severity of orthostatic hypotension, scoring from 0 to 100. |
| Orthostatic Intolerance (OI) | A clinical condition where abnormal symptoms are brought on by quiet, upright posture, often alleviated by lying down. Symptoms include abnormal heart rate increase and/or blood pressure decreases, leading to dizziness and possibly fainting. |
| Active (Orthostatic) Stand Test | An assessment measuring blood pressure and heart rate changes from lying down to standing, assessing cardiovascular responses to postural change. |
| Polymerase Chain Reaction (PCR) | A laboratory technique used to amplify DNA segments to test for the presence or absence of clinical conditions, such as infectious diseases and genetic mutations. |
| Postural Orthostatic Tachycardia Syndrome (POTS) | A clinical diagnosis where patients present with excessive increase in heart rate upon assuming upright posture or physical activity |
|  | like dizziness and palpitations. (Adult diagnostic criteria requires 30 bpm heart rate change when going from supine to upright position) |
| RAND 36-Item Health Survey (RAND-36) | A 36-item questionnaire assessing quality of life across multiple domains like physical functioning, pain, and mental health, scoring from 0-100. |
| Research Electronic Data Capture (REDCap) | A web-based application developed to support secure data capture for research studies. |
| Shapiro-Wilk Test | A statistical test used to assess the normality of a distribution within a sample data set. |
| Spearman Correlation Coefficient | A measure used to determine strength and direction of association between two ranked variables. |
| Sphygmomanometer | A device used to measure blood pressure. |
| Stimulants | Substances that increase physiological or nervous activity in the body. |
| Supine Position | Lying horizontally with the face and torso facing up. |
| Wake-Promoting Agents | Medications used to increase alertness and wakefulness. |
| Wilcoxon Rank Sum Test | A non-parametric statistical test that compares two independent samples to evaluate whether their population mean ranks differ. |

## Abbreviations

***Abbreviations-***
| **Abbreviation** | **Term** |
| --- | --- |
| COMPASS-31 | Composite Autonomic Symptom Score-31 |
| COVID-19 | Coronavirus Disease 2019 |
| EDS | Ehlers-Danlos Syndrome |
| FSS | Fatigue Severity Scale |
| GAD7 | General Anxiety Disorder-7 |
| IgG | Immunoglobulin G |
| IST | Inappropriate Sinus Tachycardia |
| LC | Long-COVID |
| LDN | Low-Dose Naltrexone |
| MCAS | Mast Cell Activation Syndrome |
| OH | Orthostatic Hypotension |
| OHQ | Orthostatic Hypotension Questionnaire |
| PASC | Post-Acute Sequelae of SARS-CoV-2 |
| PCR | Polymerase Chain Reaction |
| POTS | Postural Orthostatic Tachycardia Syndrome |
| RAND-36 | RAND 36-Item Health Survey |
| REDCap | Research Electronic Data Capture |

## Notes

### Competing Interest Statement

The authors have declared no competing interest.

### Funding Statement

This study did not receive any funding

### Author Declarations

The study received ethical approval from the Stanford University Institutional Review Board.

## References

1. Larsen NW, Stiles LE, Shaik R, et al (2022) Characterization of autonomic symptom burden in long COVID: A global survey of 2,314 adults. Front Neurol 13:. 10.3389/FNEUR.2022.1012668

2. Morrow, A.K., Malone, L.A., Kokorelis, C. et al. Long-Term COVID 19 Sequelae in Adolescents: the Overlap with Orthostatic Intolerance and ME/CFS. Curr Pediatr Rep 10, 31–44 (2022). 10.1007/s40124-022-00261-4

3. Dennis A, Cuthbertson DJ, Wootton D, et al. Multi-organ impairment and long COVID: a 1-year prospective, longitudinal cohort study. Journal of the Royal Society of Medicine. 2023;116(3):97–112. doi:10.1177/01410768231154703

4. Hastie, C.E., Lowe, D.J., McAuley, A. et al. Natural history of long-COVID in a nationwide, population cohort study. Nat Commun 14, 3504 (2023).

5. Amin-Chowdhury, Zahin, et al. Characterising Post-Covid Syndrome More than 6 Months after Acute Infection in Adults; Prospective Longitudinal Cohort Study, England, 24 Mar. 2021, 10.1101/2021.03.18.21253633.

6. Freeman R, Wieling W, Axelrod FB, Benditt DG, Benarroch E, Biaggioni I, Cheshire WP, Chelimsky T, Cortelli P, Gibbons CH, Goldstein DS, Hainsworth R, Hilz MJ, Jacob G, Kaufmann H, Jordan J, Lipsitz LA, Levine BD, Low PA, Mathias C, Raj SR, Robertson D, Sandroni P, Schatz I, Schondorff R, Stewart JM, van Dijk JG. Consensus statement on the definition of orthostatic hypotension, neurally mediated syncope and the postural tachycardia syndrome. Clin Auton Res. 2011 Apr;21(2):69–72.

7. StataCorp. 2023. Stata Statistical Software: Release 18. College Station, TX: StataCorp LLC

8. Python Software Foundation. Python Language Reference, version 3.12.1. Available at https://www.python.org/.

9. MATLAB. (2023). Version R2023b. Natick, Massachusetts: The MathWorks Inc.

10. Hernandez-Ronquillo L, Moien-Afshari F, Knox K, Britz J, Tellez-Zenteno JF. How to measure fatigue in epilepsy? The validation of three scales for clinical use. Epilepsy Res. 2011 Jun;95(1-2):119–29. doi: 10.1016/j.eplepsyres.2011.03.010. Epub 2011 Apr 8. PMID: 21482076.

11. Steward AL, Sherbourne C, Hayes RD, et al. “Summary and Discussion of MOS Measures,” in Stewart AL & Ware JE (eds.), Measuring Functioning and Well-Being: The Medical Outcome Study Approach (pp. 345–371). Durham, NC: Duke University Press, 1992.

12. Miller AJ, Stiles LE, Sheehan T, Bascom R, Levy HP, Francomano CA, Arnold AC. Prevalence of hypermobile Ehlers-Danlos syndrome in postural orthostatic tachycardia syndrome. Auton Neurosci. 2020 Mar;224:102637. doi: 10.1016/j.autneu.2020.102637. Epub 2020 Jan 10. PMID: 31954224; PMCID: PMC7282488.

13. Hira, R., Karalasingham, K., Baker, J.R. et al. Autonomic Manifestations of Long-COVID Syndrome. Curr Neurol Neurosci Rep 23, 881–892 (2023). 10.1007/s11910-023-01320-z

14. Hira, Rashmin, et al. “Objective Hemodynamic Cardiovascular Autonomic Abnormalities in Post-Acute Sequelae of COVID-19.” Canadian Journal of Cardiology, vol. 39, no. 6, June 2023, pp. 767–775, doi:10.1016/j.cjca.2022.12.002.

15. Seeley MC, Gallagher C, Ong E, Langdon A, Chieng J, Bailey D, Page A, Lim HS, Lau DH. High Incidence of Autonomic Dysfunction and Postural Orthostatic Tachycardia Syndrome in Patients with Long COVID: Implications for Management and Health Care Planning. Am J Med. 2023 Jun 29:S0002-9343(23)00402-3. doi: 10.1016/j.amjmed.2023.06.010. Epub ahead of print. PMID: 37391116; PMCID: PMC10307671.

16. Wynberg, Elke, et al. “The Effect of SARS-COV-2 Vaccination on Post-Acute Sequelae of Covid-19 (PASC): A Prospective Cohort Study.” Vaccine, vol. 40, no. 32, July 2022, pp. 4424–4431, doi:10.1016/j.vaccine.2022.05.090.

