## Supplemental Tables for "Evaluating Long-Term Autonomic Dysfunction and Functional Impacts of Long COVID: A Follow-Up Study"

**Supplementary Table 1. Logistic regression evaluating predictors of COMPASS-31 score designation.**

| **Variable** | **Odds Ratio** | **Std. Err.** | **z** | **P>\|z\|** | **[95% Conf. Interval] Lower** | **[95% Conf. Interval] Upper** |
|---|---|---|---|---|---|---|
| **Age Category** | | | | | | |
| <40 | 1.0 | | | | | reference |
| 40-44 | 1.11 | 0.42 | 0.27 | 0.78 | 0.53 | 2.34 |
| 45-49 | 0.59 | 0.22 | -1.41 | 0.16 | 0.29 | 1.23 |
| 50-54 | 0.41 | 0.14 | -2.59 | 0.01 | 0.21 | 0.81 |
| 55+ | 0.84 | 0.27 | -0.55 | 0.59 | 0.44 | 1.59 |
| **Assigned Sex at Birth** | | | | | | |
| Female | 1.0 | | | | | reference |
| Male | 0.44 | 0.14 | -2.63 | 0.01 | 0.24 | 0.81 |
| **Ethnicity** | | | | | | |
| Hispanic/Latino | 1.0 | | | | | reference |
| Not Hispanic | 1.07 | 0.52 | 0.13 | 0.90 | 0.41 | 2.77 |
| Prefer not to disclose | 0.67 | 0.58 | -0.46 | 0.64 | 0.12 | 3.71 |
| Unknown | 0.91 | 1.37 | -0.06 | 0.95 | 0.05 | 17.30 |
| **Vaccination Status** | | | | | | |
| Unvaccinated | 1.0 | | | | | reference |
| Vaccinated | 0.39 | 0.26 | -1.43 | 0.152 | 0.11 | 1.42 |
| **Country** | | | | | | |
| U.S. Residence | 1.0 | | | | | reference |
| Outside U.S. | 0.33 | 0.08 | -4.51 | 0.0 | 0.20 | 0.53 |
| **Autoimmune Dx** | | | | | | |
| No | 1.0 | | | | | reference |
| yes | 1.50 | 0.53 | 1.21 | 0.23 | 0.78 | 2.90 |
| **Joint Hypermobility** | | | | | | |
| No | 1.0 | | | | | reference |
| Yes | 1.68 | 0.43 | 2.01 | 0.04 | 1.013 | 2.79 |
| **_cons** | 12.42 | 7.79 | 4.01 | 0.0 | 3.63 | 42.49 |

### Supplementary Table 2. Post-COVID POTS diagnosis logistic regression

| Variable | Odds Ratio | Std. Err. | z | P>\|z\| | [95% Conf. Interval] Lower | [95% Conf. Interval] Upper |
|---|---|---|---|---|---|---|
| **Age Category** | | | | | | |
| <40 | 1.0 | | | | | reference |
| 40-44 | 0.88 | 0.28 | -0.39 | 0.70 | 0.48 | 1.64 |
| 45-49 | 0.55 | 0.19 | -1.76 | 0.08 | 0.28 | 1.07 |
| 50-54 | 0.36 | 0.12 | -3.13 | 0.002 | 0.19 | 0.68 |
| 55+ | 0.51 | 0.15 | -2.36 | 0.018 | 0.29 | 0.89 |
| **Assigned Sex at Birth** | | | | | | |
| Female | 1.0 | | | | | reference |
| Male | 0.64 | 0.22 | -1.29 | 0.20 | 0.33 | 1.26 |
| **Ethnicity** | | | | | | |
| Hispanic/Latino | 1.0 | | | | | reference |
| Not Hispanic | 0.92 | 0.40 | -0.19 | 0.85 | 0.39 | 2.15 |
| Prefer not to disclose | 0.47 | 0.44 | -0.81 | 0.42 | 0.07 | 2.92 |
| Unknown[1] | | | | | | |
| **Vaccination Status** | | | | | | |
| Unvaccinated | 1.0 | | | | | reference |
| Vaccinated | 0.40 | 0.32 | -1.14 | 0.25 | 0.08 | 1.94 |
| **Country** | | | | | | |
| U.S. Residence | 1.0 | | | | | reference |
| Outside U.S. | 0.82 | 0.18 | -0.93 | 0.35 | 0.53 | 1.25 |
| **Autoimmune Dx** | | | | | | |
| No | 1.0 | | | | | reference |
| yes | 1.48 | 0.42 | 1.38 | 0.17 | 0.85 | 2.59 |
| **Joint Hypermobility** | | | | | | |
| No | 1.0 | | | | | reference |
| Yes | 2.06 | 0.45 | 3.33 | 0.001 | 1.35 | 3.17 |
| **_cons** | 0.95 | 0.52 | -0.09 | 0.93 | 0.33 | 2.76 |

[1] Not enough participant information represented for this specific logistic regression analysis

### Supplementary Table 3. RAND-36 LC cohort versus population Scores

Assessment of Rand-36 instruments reliability coefficients and comparison between population norms[11] and the PASC cohort means. Crohnbach's Alpha reliability coefficients imply that the scales effectively measure their intended instruments*.

| Scale | Items | Alpha* | RAND Corporation (N=2,471), Mean (SD) | Survey Participants (N=491), Mean (SD) |
|---|---|---|---|---|
| **Physical functioning** | 10 | 0.93 | 70.6 (27.4) | 50.6 (29.5) |
| **Role functioning/physical** | 4 | 0.84 | 53.0 (40.8) | 22.1 (3.8) |
| **Role functioning/emotional** | 3 | 0.83 | 65.8 (40.2) | 54.2 (45.2) |
| **Energy/fatigue** | 4 | 0.86 | 52.2 (22.4) | 31.5 (21.9) |
| **Emotional well-being** | 5 | 0.89 | 70.4 (22.0) | 65.0 (19.7) |
| **Social functioning** | 2 | 0.85 | 78.8 (25.4) | 44.9 (30.5) |
| **Pain** | 2 | 0.78 | 77.1 (25.5) | 53.4 (26.8) |
| **General health** | 5 | 0.78 | 57.0 (21.1) | 35.9 (21.4) |

### Supplementary Table 4. RAND-36 cohort scores characterized by COMPASS-31 score group

Comparative analysis of health-related quality of life measures between survey participants with COMPASS-31 scores <20 and ≥20: mean and median values with statistical significance.

| Scale | Items | COMPASS-31<20 Mean (SD) | COMPASS-31<20 Median [IQR] | COMPASS-31≥20 Mean (SD) | COMPASS-31≥20 Median [IQR] | FDR p-Value |
|---|---|---|---|---|---|---|
| **Physical function** | 10 | 68.9 (25.0) | 75 [50-91] | 44.3 (29.2) | 40 [22.2-70] | <0.001 |
| **Physical Limitations** | 4 | 41.7 (43.2) | 25 [0-100] | 14.7 (27.8) | 0 [0-25] | <0.001 |
| **Emotional limitations** | 3 | 63.2 (43.3) | 100 [0-100] | 50.8 (45.4) | 66.7 [0-100] | 0.005 |
| **Energy/Fatigue** | 4 | 44.2 (22.3) | 39 [27.5-59] | 26.8 (19.7) | 26 [11-39] | <0.001 |
| **Wellbeing** | 5 | 69.9 (18.0) | 76 [56-84] | 63.2 (20.1) | 64 [52-80] | <0.001 |
| **Social Functioning** | 2 | 62.2 (30.6) | 62.5 [37.5-87.5] | 38.4 (27.8) | 37.5 [12.5-62.5] | <0.001 |
| **Pain** | 2 | 70.5 (25.0) | 77.5 [45-90] | 47.0 (24.6) | 45 [32.5-67.5] | <0.001 |
| **General Health** | 5 | 46.9 (20.6) | 45 [30-61.5] | 31.7 (20.2) | 27.5 [15-45] | <0.001 |

### Supplementary Table 5. Orthostatic Standing Test Results (N=86)

| Measure | Timepoint | All OI Complaints (N=86) | COMPASS-31 >= 20 (N=68) | COMPASS-31 < 20 (N=18) | COMPASS-31 p-Value | POTS Criteria Met (N=33) | No POTS Criteria (N=53) | POTS Criteria p-Value |
|---|---|---|---|---|---|---|---|---|
| **Heart Rate (bpm):** median [IQR] | Baseline | 70 [62-74] | 70 [61-74] | 70 [67-75] | 0.82 | 72 [59-77] | 69 [62-74] | 0.69 |
| | Max | 91.5 [83-109] | 92 [83.5-115.5] | 89 [83-95] | 0.12 | 115 [104-130] | 85 [75-92] | p<0.001 |
| | Delta | 22 [14-38] | 27 [15.5-39] | 17 [12-28] | 0.07 | 40 [36-52.5] | 16 [11-20] | p<0.001 |
| **Systolic Blood Pressure (mmHg):** median [IQR] | Baseline | 115.5 [104-125] | 113.5 [104-125.5] | 121.5[107-125] | 0.46 | 115 [105-130.5] | 117 [104-125] | 0.90 |
| | Max | 130 [120-140] | 130 [119.5-138] | 130.5 [120-144] | 0.61 | 132 [124-144] | 129 [118-137] | 0.38 |
| | Delta | 13 [5-22] | 12.5 [5.5-21.5 | 15 [5-22] | p>0.99 | 15 [3-25.5] | 12 [7-18] | 0.49 |
| **Diastolic Blood Pressure (mmHg):** median [IQR] | Baseline | 69.5 [64-80] | 69.5 [62.5-77.5] | 70 [66-84] | 0.31 | 72 [64-80] | 69 [64-77] | 0.58 |
| | Max | 88.5 [80-94] | 87 [79.5-93.5] | 89 [83-94] | 0.49 | 90 [83-98] | 86 [80-93] | 0.10 |
| | Delta | 15 [9-23] | 15 [10-22.5] | 17.5 [7-24] | p>0.99 | 20 [9.5-25] | 14 [9-19] | 0.21 |
